# COVID 19: Real-time Forecasts of confirmed cases, active cases, and health infrastructure requirements for India and its states using the ARIMA model

**DOI:** 10.1101/2020.05.17.20104588

**Authors:** Rishabh Tyagi, Mahadev Bramhankar, Mohit Pandey, M Kishore

**Author notes:** Co-first authors (Rishabh Tyagi, Mahadev Bramhankar).

## Abstract

**Background:** COVID-19 is an emerging infectious disease which has been declared a Pandemic by the World Health Organization (WHO) on 11th March 2020. The Indian public health care system is already overstretched, and this pandemic is making things even worse. That is why forecasting cases for India is necessary to meet the future demands of the health infrastructure caused due to COVID-19.

**Objective:** Our study forecasts the confirmed and active cases for COVID-19 until July mid, using time series Autoregressive Integrated Moving Average (ARIMA) model. Additionally, we estimated the number of isolation beds, Intensive Care Unit (ICU) beds and ventilators required for the growing number of COVID-19 patients.

**Methods:** We used ARIMA model for forecasting confirmed and active cases till the 15th July. We used time-series data of COVID-19 cases in India from 14th March to 22nd May. We estimated the requirements for ICU beds as 10%, ventilators as 5% and isolation beds as 85% of the active cases forecasted using the ARIMA model.

**Results:** Our forecasts indicate that India will have an estimated 7,47,772 confirmed cases (95% CI: 493943, 1001601) and 296,472 active cases (95% CI:196820, 396125) by 15th July. While Maharashtra will be the most affected state, having the highest number of active and confirmed cases, Punjab is expected to have an estimated 115 active cases by 15th July. India needs to prepare 2,52,001 isolation beds (95% CI: 167297, 336706), 29,647 ICU beds (95% CI: 19682, 39612), and 14,824 ventilator beds (95% CI: 9841, 19806).

**Conclusion:** Our forecasts show an alarming situation for India, and Maharashtra in particular. The actual numbers can go higher than our estimated numbers as India has a limited testing facility and coverage.

## 1. Introduction

COVID-19 is an emerging infectious disease caused by severe acute respiratory syndrome Corona Virus 2 (SARS-CoV-2). The first human cases of COVID-19 were first reported by officials in Wuhan City, China, in December 2019. Retrospective investigations by Chinese authorities have identified human cases with onset of symptoms in early December 2019. While some of the earliest known cases had a link to a wholesale food market in Wuhan, some did not (WHO, 2020). The World Health Organization (WHO) has declared the outbreak of the novel Coronavirus (COVID-19) as a pandemic on 11th March 2020.

This pandemic is spreading very quickly throughout the world, and the number of confirmed cases reached 6.44 million on 2nd June. Globally, the recovery rate is around 46.7 percent and case fatality rate of 5.9 percent (Worldometer, 2nd June 2020). On 2nd June, India’s confirmed cases of COVID-19 crossed 2 Lakhs. The recovery rate was 48.31 percent amongst COVID-19 patients. The case fatality rate of COVID-19 in India declined to 2.8 percent on 2nd June in India. They also told that 73 percent of COVID-19 deaths in India are people with co-morbidities (Press Information Bureau 2020 c).

In India, the first positive case of COVID-19 was detected on 30th January 2020, in Kerala (India Today, 2020). With the experience of the overwhelming burden of COVID-19 in Europe and China, the Government of India implemented a complete nationwide lockdown on 25th March for 21 days (Press Information Bureau 2020 a). The purpose of the nationwide lockdown was to contain the spread of the Coronavirus so that the Government could take a multi-prong strategy: add more beds in its network of hospitals, scale up the production of the testing kits for COVID-19 and personal protection equipment (PPE) for the health workers. On 3rd June, with respect to the health infrastructure in the country for the management of COVID-19, 952 dedicated COVID hospitals with 1,66,332 Isolation beds, 21,393 ICU beds, and 72,762 Oxygen supported beds are available. 2,391 dedicated COVID Health Centres with 1,34,945 Isolation beds; 11,027 ICU beds and 46,875 Oxygen supported beds have been operationalized. (Press Information Bureau 2020 c).

A few studies have attempted to predict numbers of confirmed and active cases at the national and state level for India to assess the burden of COVID-19 in future. Tiwari et al. (2020) made their prediction for India based on the pattern of China using a machine learning approa0ch.

They predicted that the peak of the cases for India would be attained between the third and fourth weeks of April 2020 in India. The infection was likely to be controlled by the end of May 2020. Chakraborty (2020) used a hybrid approach, based on an ARIMA and Wavelet-based forecasting model, to make short-term forecasts of the number of daily confirmed cases in Canada, France, India, South Korea, and the United Kingdom. The ARIMA model has been used in many studies for forecasting case count of epidemic diseases based on the time series modelling (Gupta and Pal 2020; Tandon et al., 2020; Kumar et al. 2020; Perone, 2020).

In India, the healthcare infrastructure is not up to the mark. The number of hospital beds per 1000 population is less than one - it is just one indicator to cite the vulnerable situation of India’s health care systems (World Bank Database). Several studies from countries like China, Italy and Spain show the requirements of different healthcare facilities required to fight this pandemic. Guan et al. (2020) in his study on data regarding 1099 patients with laboratory-confirmed COVID-19 from 552 hospitals in 30 provinces, in mainland China through 29th January 2020, found that 5.0% were admitted to the ICU, 2.3% who underwent invasive mechanical ventilation, and 1.4% who died. A study by Lazzerini et al. (2020) for Italy and Spain suggested that 40-55% of COVID-19 positive cases have been hospitalized, with 7-12% requiring admission to intensive care units. The COVID-19 Cases in Italy shows that 10-25% of patients will require ventilation, and some patients will need ventilation for several weeks. Remuzzi et al., (2020) find that the percentage of patients in intensive care reported daily in Italy between 1st March and 11th March 2020, has consistently been between 9% and 11% of patients who are actively infected.

In the Indian scenario, a press release by PIB on 8th May stated that of the total 35,902 active cases, 4.8 per cent patients are in ICU, 1.1 per cent on ventilators and 3.3 per cent are on oxygen support (Press Information Bureau 2020 b). With the rising number of cases, it is crucial to forecast the requirements of healthcare infrastructure like isolation beds, ICU beds and ventilators at national and state level to make the respective authorities aware of the situation they might be facing shortly. In this study, we use the well-known ARIMA time-series model to forecast the confirmed and active cases for India at national and state level. Another objective of this study is to estimate the requirements of healthcare infrastructure in the future based on the forecasts of active cases.

## 2. Data and Methods

### 2.1 Data

Data on COVID-19 was obtained from the data-sharing portal covid19india.org. Information is collected on daily confirmed and active cases at the national and state level from 14^th^ March to 22^nd^ May 2020. This dataset provides excel of the patient database, which is used to build a required time-series. From this dataset, we have used cumulative confirmed and active cases for India and selected states. Selection of states is based on the criterion that states should have at least 100 confirmed cases on 3rd May 2020. By using this selection criterion, India and 17 other states selected which are Andhra Pradesh, Bihar, Delhi, Gujarat, Haryana, Jammu & Kashmir, Odisha, Karnataka, Kerala, Madhya Pradesh, Maharashtra, Punjab, Rajasthan, Tamil Nadu, Telangana, Uttar Pradesh, West Bengal.

### 2.2 Method

In this study, we applied the ARIMA model to our considered time series data of COVID-19 cases by using R-studio, for forecasting cumulative confirmed and active cases. This model has been preferred for the time series forecasting in various fields as the model predictions are based on different parameters. The required parameters for the ARIMA model are *(p, d, q)* which evaluate autoregressive term, integrated moving average and past lag term for stationary time series respectively. The degree of parameters p, d and q are determined based on the partial Auto-correlation function (PACF), Augmented Dickey-Fuller Test to test the stationary time series and Complete Auto-Correlation Function (ACF) respectively. These parameters help to capture overall fluctuations in earlier time-series which helps to predict the future. ARIMA has some advantages over the other models as it not only captures the overall picture in earlier trend, but it also provides a 95% confidence interval (CI) for our point estimates.

The model for forecasting future confirmed and active cases of COVID-19 cases is represented as,

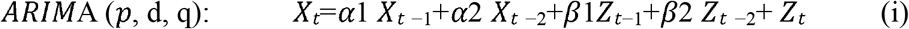

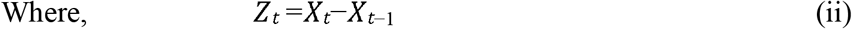

Here, *X_t_* is the predicted number of cumulative confirmed and active COVID-19 cases at *t^h^* day; *α*2, *α*2, *β*1 and *β*2 are parameters whereas *Z_t_* is the residual term for *t^th^* day.

Finally, we estimate the requirement of health infrastructure, i.e., the requirement of Intensive care unit (ICU), Ventilator support, and isolation beds using the following equations based on forecasted active cases.

**Required number of beds** = Forecasted active cases at ith day*(85/100)

**Required number of ICU** = Forecasted active cases at ith day*(10/100)

**Required number of Ventilators** = Forecasted active cases at ith day*(5/100)

Based on these equations, infrastructure estimates till the mid of July.

## 3. Results

### 3.1 ARIMA model fit for confirmed and active cases of COVID-19 in India

Fig 1(a) & 1(b), shows the ARIMA model fitted correlogram for the active and confirmed cases. In these figures, we see four subfigures which reveal the trend for the earlier and forecasted values for both confirmed and active cases. Forecasting based on PACF and ACF graphs helps to determine parameters p and q. Moreover, the best ARIMA model fit is considered having the lowest Akaike Information Criterion (AIC) value.

**Fig. 1(a).**
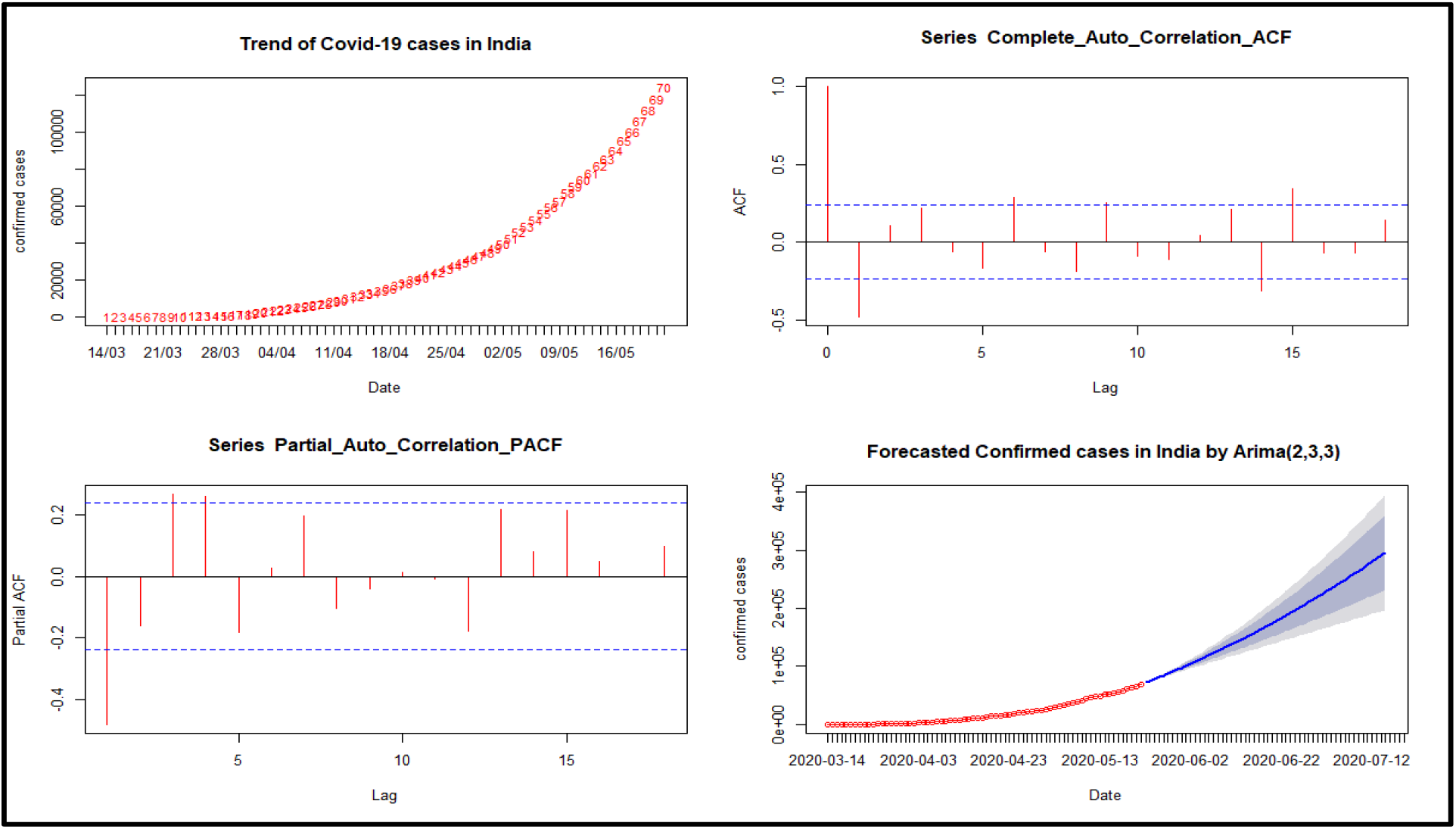
Correlogram and ARIMA forecast for the Confirmed COVID-19 Cases in India.

**Fig. 1(b).**
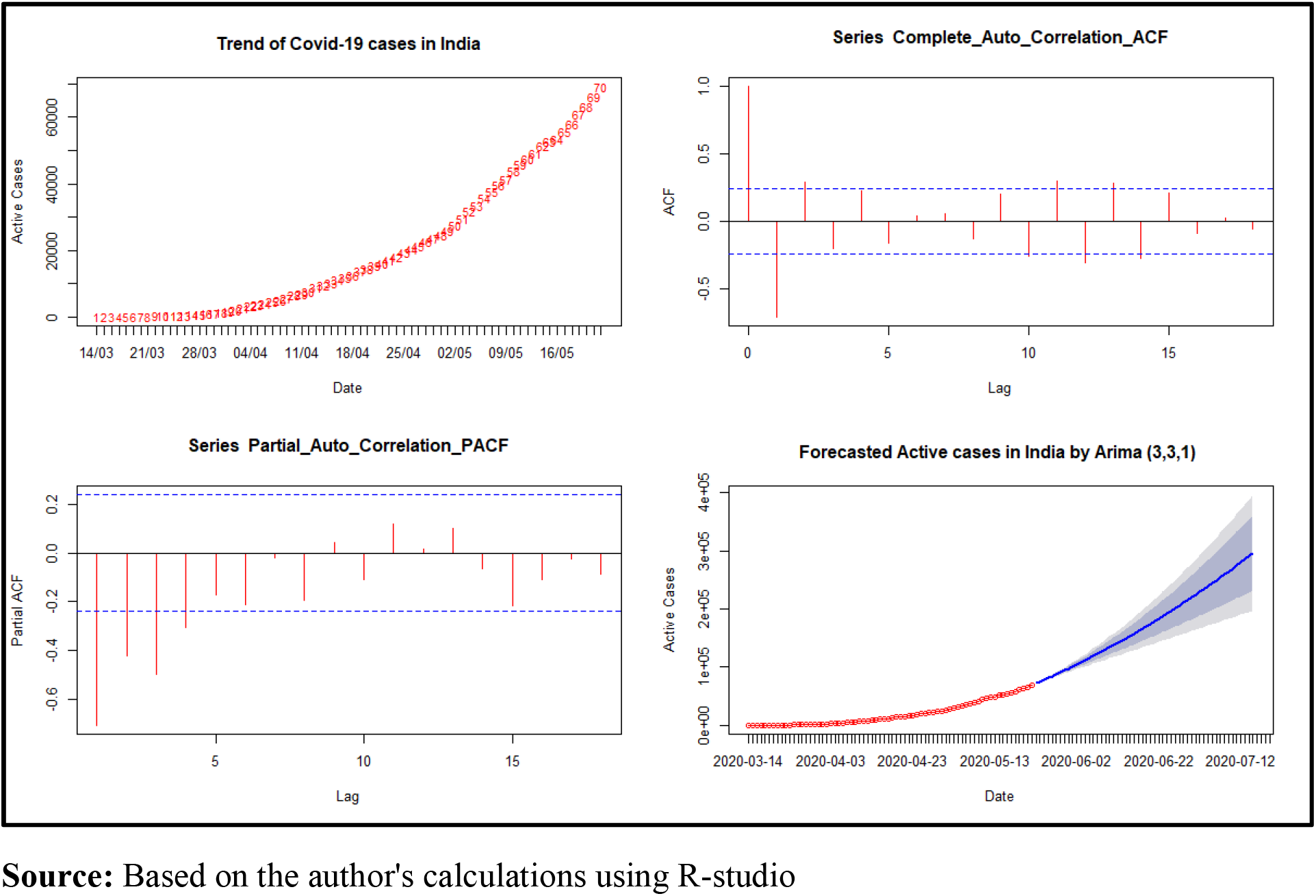
Correlogram and ARIMA Forecast for the Active COVID-19 Cases in India.

Fig 1(a) also shows the fitted model for total confirmed cases with ARIMA (2, 3, 3) having low AIC. With the help of this model, we have predicted future confirmed cases with 95% CI till 15th July 2020. Similarly, for total active cases, we have a suitable model with ARIMA (3, 3, 1) which helps to predict active cases for the same duration.

### 3.2 Forecasted Confirmed and active cases of COVID-19 for India

The analysis result for India in **Table 1 and Fig. 2** shows the total confirmed and active cases for India. It has observed that with time, confirmed cases will increase at a faster rate. India will be having 5,19,258 confirmed cases (95% CI: 389542, 648794) at June 30 and 7,47,772 confirmed cases (95% CI: 493943, 1001601) till July 15.

**Table 1:** Forecast of Confirmed and Active Cases of COVID-19 for India from 23rd May to 15th July, 2020,

| Date | Confirmed Cases | 95% Confidence Interval |  | Active Cases | 95% Confidence Interval |  | Death & Recovered (%) |
| --- | --- | --- | --- | --- | --- | --- | --- |
|  | Point Estimates | Lower | Upper | Point Estimates | Lower | Upper |  |
| 23 May 2020 | 131258 | 130705 | 131810 | 73186 | 71515 | 72858 | 44 |
| 25 May 2020 | 144843 | 143116 | 146570 | 78244 | 76558 | 79930 | 46 |
| 27 May 2020 | 159100 | 155473 | 162727 | 84551 | 81574 | 87528 | 47 |
| 29 May 2020 | 174321 | 168258 | 180384 | 91021 | 86476 | 95566 | 48 |
| 31 May 2020 | 190164 | 181148 | 199179 | 97682 | 91332 | 104032 | 49 |
| 02 June 2020 | 206764 | 194194 | 219334 | 104526 | 96145 | 112907 | 49 |
| 04 June 2020 | 224206 | 207521 | 240891 | 111556 | 100930 | 122182 | 50 |
| 06 June 2020 | 242329 | 220948 | 263711 | 118771 | 105691 | 131851 | 51 |
| 08 June 2020 | 261231 | 234534 | 287928 | 126171 | 110433 | 141909 | 52 |
| 10 June 2020 | 280918 | 248292 | 313545 | 133756 | 115161 | 152351 | 52 |
| 12 June 2020 | 301326 | 262133 | 340520 | 141527 | 119877 | 163176 | 53 |
| 14 June 2020 | 322510 | 276089 | 368930 | 149482 | 124584 | 174380 | 54 |
| 16 June 2020 | 344458 | 290144 | 398772 | 157623 | 129282 | 185963 | 54 |
| 18 June 2020 | 367148 | 304251 | 430044 | 165949 | 133974 | 197924 | 55 |
| 20 June 2020 | 390607 | 318423 | 462791 | 174460 | 138659 | 210260 | 55 |
| 22 June 2020 | 414824 | 332635 | 497013 | 183156 | 143340 | 222972 | 56 |
| 24 June 2020 | 439792 | 346862 | 532722 | 192037 | 148015 | 236060 | 56 |
| 26 June 2020 | 465524 | 361103 | 569946 | 201104 | 152686 | 249522 | 57 |
| 28 June 2020 | 492013 | 375335 | 608692 | 210356 | 157352 | 263359 | 57 |
| 30 June 2020 | 519258 | 389542 | 648974 | 219793 | 162014 | 277571 | 58 |
| 02 July 2020 | 547264 | 403715 | 690814 | 229415 | 166672 | 292158 | 58 |
| 04 July 2020 | 576027 | 417835 | 734219 | 239222 | 171325 | 307119 | 58 |
| 06 July 2020 | 605547 | 431888 | 779206 | 249215 | 175973 | 322456 | 59 |
| 08 July 2020 | 635827 | 445863 | 825790 | 259392 | 180616 | 338169 | 59 |
| 10 July 2020 | 666863 | 459742 | 873984 | 269755 | 185253 | 354257 | 60 |
| 12 July 2020 | 698658 | 473515 | 923801 | 280303 | 189884 | 370722 | 60 |
| 14 July 2020 | 731212 | 487167 | 975256 | 291036 | 194509 | 387563 | 60 |
| 15 July 2020 | 747772 | 493943 | 1001601 | 296472 | 196820 | 396125 | 60 |

**Fig. 2.**
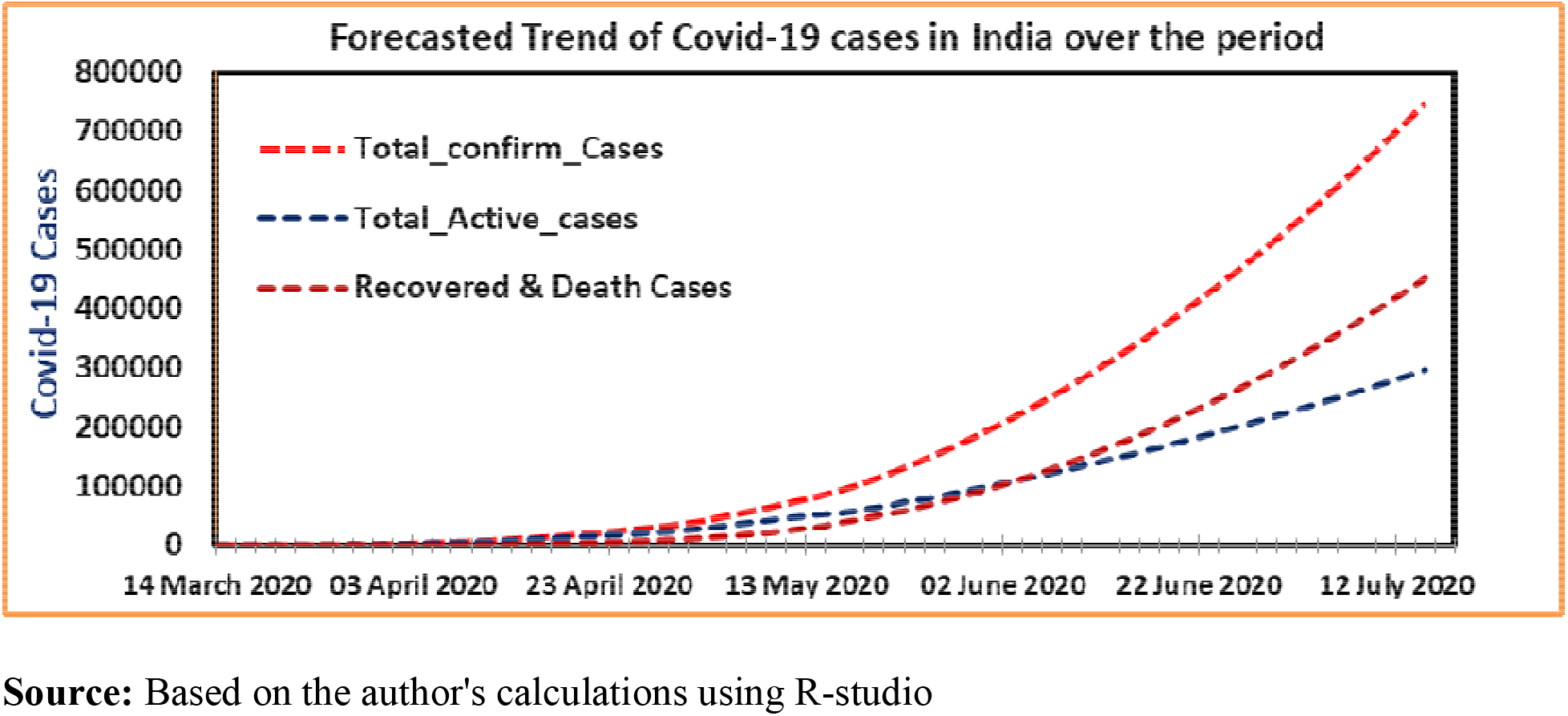
Trend of Forecasted Confirm and Active COVID-19 Cases over the Period in India.

Similarly, based on our forecasts for active cases for India, we found that active cases will be 2,19,793 (95% CI: 162014, 277571) at 30th June and 2,96,472 active cases (95% CI: 196820, 396125) at 15th July. The combined number of recovered and death cases are also increasing rapidly and are expected to be around 60 percent of total confirmed cases by 15th July based on our forecasts.

### 3.3 Forecasted Confirmed cases of COVID-19 for the selected states of India

**Table 2**, shows the forecast of confirmed for selected states of India on 15th June, 30th June and 15th July. Our forecasts show that that confirmed cases for states vary a lot from one state to another. Our forecasts for 15th June for confirmed cases show that Haryana, Punjab and Kerala will have around 2000 confirmed cases. However, states like Maharashtra 108712 confirmed cases (95% CI: 89631, 127793), Tamil Nadu 36759 confirmed cases (95% CI: 26703, 46816), Delhi 26257 confirmed cases (CI: 20794, 31720) & Gujarat 22860 confirmed cases (95% CI: 18187, 27533) would have a high number of confirmed cases at the 15th June. On 30th June, only Haryana and Punjab will be having around 2000 confirmed cases based on our forecasts. Though, states like Maharashtra 149072 confirmed cases (95% CI: 109237, 188908), Tamil Nadu 54263 confirmed cases (95% CI: 31834, 76692), Delhi 34968 confirmed cases (95% CI: 24090, 45847) & Gujarat 28852 confirmed cases (95% CI: 19770, 37934) will have to occur confirmed cases on 30th June. On 15th July, Punjab will be having the least number of confirmed cases around 4000 confirmed cases. However, other states like Maharashtra 189433 confirmed cases (95% CI: 124301, 254565), Tamil Nadu 74637 confirmed cases (95% CI: 35282, 113991), Delhi (43680 confirmed cases (95% CI: 26271, 61089) & Bihar 38027 confirmed cases (95% CI: 14862, 61192) will have a high number of confirmed cases at the 15th July.

**Table 2:**
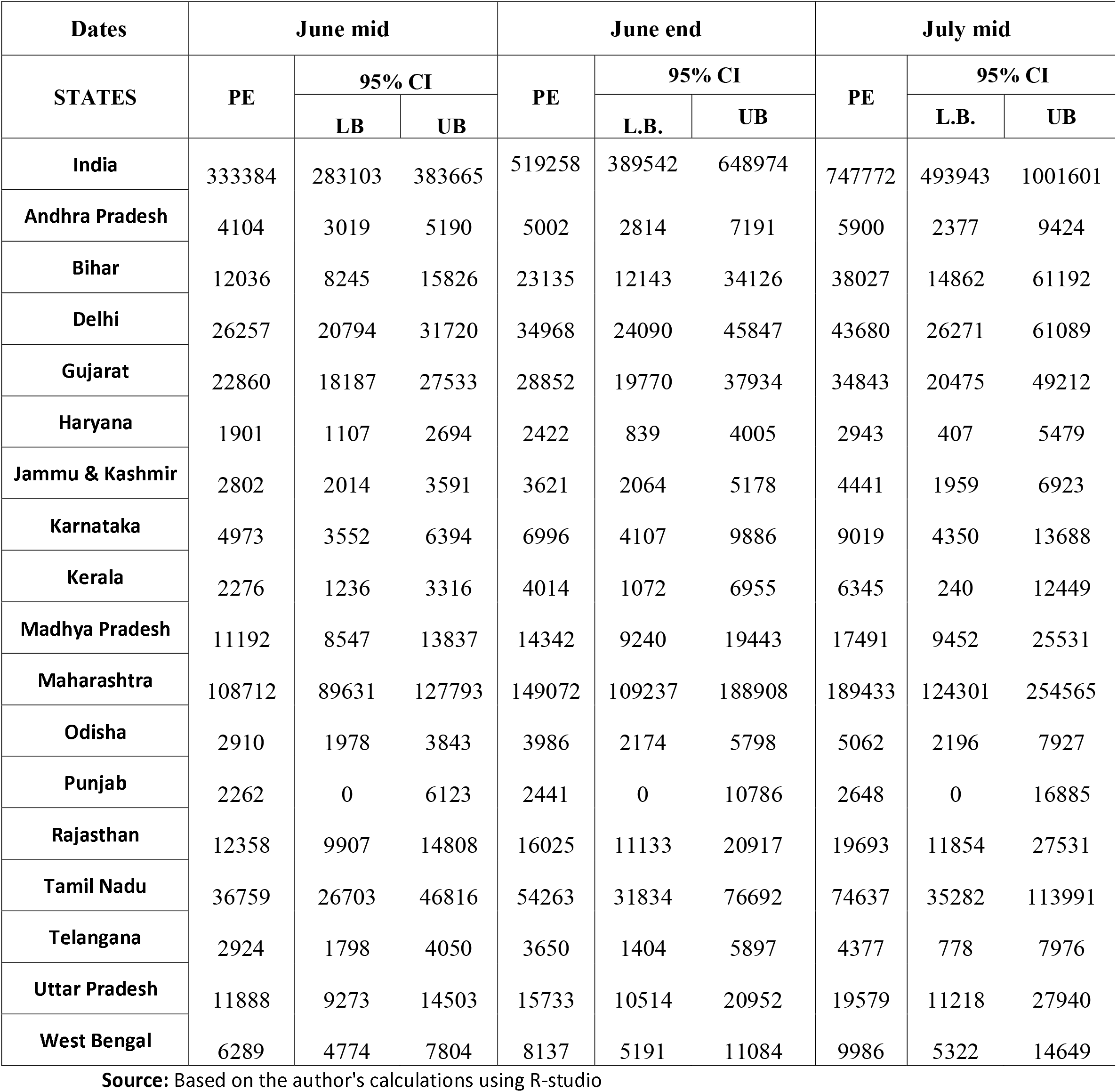
Forecast of Confirmed Cases of COVID-19 for India and its States on 15th June, 30th June and 15th July, 2020, India

### 3.4 Forecasted active cases of COVID-19 for the selected states of India

**Table 3** shows the state-wise active cases for India. On 15th June, Punjab, Kerala and Andhra Pradesh will be having less than 1000 active cases based on our forecasts. However, states like Maharashtra (67984 active cases (95% CI: 56127, 79840)), Delhi (10907 active cases (95% CI: 5590, 16224)) & Tamil Nadu (8534 active cases (95% CI: 0, 21421)) will have a high number of active cases on the 15th June.

**Table 3:** Forecast of Active Cases of COVID-19 for India and its States on 15th June, 30th June and 15th July, 2020, India

| Dates | June mid |  |  | June end |  |  | July mid |  |  |
| --- | --- | --- | --- | --- | --- | --- | --- | --- | --- |
| STATES | PE | 95% CI |  | PE | 95% CI |  | PE | 95% CI |  |
|  |  | LB | UB |  | L.B. | UB |  | L.B. | UB |
| India | 153529 | 126934 | 180125 | 219793 | 162014 | 277571 | 296472 | 196820 | 396125 |
| Andhra Pradesh | 1000 | 0 | 2180 | 1065 | 0 | 3327 | 1131 | 0 | 4685 |
| Bihar | 6922 | 3684 | 10160 | 11680 | 3864 | 19496 | 17479 | 2997 | 31961 |
| Delhi | 10907 | 5590 | 16224 | 13840 | 3386 | 24294 | 16773 | 142 | 33404 |
| Gujarat | 9133 | 5175 | 13091 | 10716 | 3213 | 18219 | 12299 | 572 | 24027 |
| Haryana | 1057 | 0 | 2470 | 1589 | 0 | 4618 | 2200 | 0 | 7351 |
| Jammu & Kashmir | 1758 | 1079 | 2437 | 2372 | 1040 | 3705 | 2986 | 867 | 5105 |
| Karnataka | 4203 | 2579 | 5826 | 6121 | 2820 | 9422 | 8039 | 2707 | 13372 |
| Kerala | 918 | 102 | 1734 | 1362 | 0 | 3035 | 1807 | 0 | 4520 |
| Madhya Pradesh | 5109 | 1629 | 8589 | 6487 | 0 | 13379 | 7864 | 0 | 18866 |
| Maharashtra | 67984 | 56127 | 79840 | 91428 | 67487 | 115369 | 114873 | 76308 | 153438 |
| Odisha | 1285 | 330 | 2241 | 1627 | 0 | 3573 | 1969 | 0 | 5116 |
| Punjab | 115 | 0 | 2305 | 115 | 0 | 2967 | 115 | 0 | 3502 |
| Rajasthan | 6586 | 3884 | 9288 | 8962 | 3571 | 14353 | 11338 | 2703 | 19972 |
| Tamil Nadu | 8534 | 0 | 21421 | 9163 | 0 | 35097 | 9793 | 0 | 51497 |
| Telangana | 1399 | 100 | 2697 | 1854 | 0 | 4393 | 2310 | 0 | 6339 |
| Uttar Pradesh | 5006 | 2093 | 7919 | 6716 | 936 | 12496 | 8426 | 0 | 17660 |
| West Bengal | 3274 | 2298 | 4250 | 4157 | 2361 | 5954 | 5041 | 2275 | 7807 |

On 30th June, Haryana, Punjab, Andhra Pradesh and Kerala will be having fewer than 2000 active cases based on our forecasts. However, states like Maharashtra (91428 active cases (95% CI: 67487, 115639), Delhi (13840 active cases (95% CI: 3386, 24294) & Bihar (11680 active cases (95% CI: 3864, 19496) will have a high number of active cases on the 30th June. On 15th July, Punjab, Andhra Pradesh and Kerala will be having under 2000 active cases based on our forecasts. However, states like Maharashtra (114873 active cases (95% CI: 76308, 153438), Bihar (17479 active cases (95% CI: 2997, 31961) & Delhi (16773 active cases (95% CI: 142, 33404) will have a high number of active cases on 15th July.

### 3.5 Estimates of required healthcare infrastructure for India

**Table 4, 5, and 6** show the required number of isolation beds, ICU beds and ventilators for active cases of COVID-19 on 15th June, 30th June and 15th July, respectively for India. The total number of isolation beds required is 130500 (95% CI: 107894, 153106). It also shows that 15353 ICU beds (95% CI: 12693, 18012) and 7676 ventilators (95% CI: 6347, 9006) will be required in India by 15th June. On 30th June. The total number of isolation beds required for India is 186824 (95% CI: 137712, 235935). It also shows that 21979 ICU beds (95% CI: 16201, 27757) and 10990 ventilators (95% CI: 8101, 13879) will be required in India by 30th June. India will require 252001 isolation beds (95% CI: 167297, 336706), 29647 ICU beds (95% CI: 19682, 39612) and 14824 ventilators (95% CI: 9841, 19806) on July 15.

**Table 4:** Forecast of Active Cases of COVID-19 and Required Health Infrastructure for India and its States on 15th June, 2020, India

| STATES | Active Cases | 95% CI |  | ICU | 95% CI |  | Ventilators | 95% CI |  | Beds | 95% CI |  |
| --- | --- | --- | --- | --- | --- | --- | --- | --- | --- | --- | --- | --- |
|  | PE | LB | UB | PE | LB | UB | PE | LB | UB | PE | LB | UB |
| India | 153529 | 126934 | 180125 | 15353 | 12693 | 18012 | 7676 | 6347 | 9006 | 130500 | 107894 | 153106 |
| Andhra Pradesh | 1000 | 0 | 2180 | 100 | 0 | 218 | 50 | 0 | 109 | 850 | 0 | 1853 |
| Bihar | 6922 | 3684 | 10160 | 692 | 368 | 1016 | 346 | 184 | 508 | 5884 | 3131 | 8636 |
| Delhi | 10907 | 5590 | 16224 | 1091 | 559 | 475 | 545 | 279 | 238 | 9271 | 4751 | 4039 |
| Gujarat | 9133 | 5175 | 13091 | 913 | 518 | 1309 | 457 | 259 | 655 | 7763 | 4399 | 11127 |
| Haryana | 1057 | 0 | 2470 | 106 | 0 | 247 | 53 | 0 | 124 | 899 | 0 | 2100 |
| Jammu & Kashmir | 1758 | 1079 | 2437 | 176 | 108 | 244 | 88 | 54 | 122 | 1494 | 918 | 2071 |
| Karnataka | 4203 | 2579 | 5826 | 420 | 258 | 583 | 210 | 129 | 291 | 3572 | 2192 | 4952 |
| Kerala | 918 | 102 | 1734 | 92 | 10 | 173 | 46 | 5 | 87 | 780 | 87 | 1474 |
| Madhya Pradesh | 5109 | 1629 | 8589 | 511 | 163 | 859 | 255 | 81 | 429 | 4343 | 1385 | 7301 |
| Maharashtra | 67984 | 56127 | 79840 | 6798 | 5613 | 7984 | 3399 | 2806 | 3992 | 57786 | 47708 | 67864 |
| Odisha | 1285 | 330 | 2241 | 129 | 33 | 224 | 64 | 16 | 112 | 1093 | 280 | 1905 |
| Punjab | 115 | 0 | 2305 | 12 | 0 | 231 | 6 | 0 | 115 | 98 | 0 | 1959 |
| Rajasthan | 6586 | 3884 | 9288 | 659 | 388 | 929 | 329 | 194 | 464 | 5598 | 3302 | 7895 |
| Tamil Nadu | 8534 | 0 | 21421 | 853 | 0 | 214 | 427 | 0 | 1071 | 7254 | 0 | 18208 |
| Telangana | 1399 | 100 | 2697 | 140 | 10 | 270 | 70 | 5 | 135 | 1189 | 85 | 2293 |
| Uttar Pradesh | 5006 | 2093 | 7919 | 501 | 209 | 792 | 250 | 105 | 396 | 4255 | 1779 | 6731 |
| West Bengal | 3274 | 2298 | 4250 | 327 | 230 | 425 | 164 | 115 | 213 | 2783 | 1953 | 3613 |

**Source:** Based on the author's calculations using R-studio

**Table 5:** Forecast of Active Cases of COVID-19 and Required Health Infrastructure for India and its States on 30th June, 2020, India

| STATES | Active Cases | 95% CI |  | ICU | 95% CI |  | Ventilators | 95% CI |  | Beds | 95% CI |  |
| --- | --- | --- | --- | --- | --- | --- | --- | --- | --- | --- | --- | --- |
|  | PE | LB | UB | PE | LB | UB | PE | LB | UB | PE | LB | UB |
| India | 219793 | 162014 | 277571 | 21979 | 16201 | 27757 | 10990 | 8101 | 13879 | 186824 | 137712 | 235935 |
| Andhra Pradesh | 1065 | 0 | 3327 | 107 | 0 | 333 | 53 | 0 | 166 | 906 | 0 | 2828 |
| Bihar | 11680 | 3864 | 19496 | 1168 | 386 | 1950 | 584 | 193 | 975 | 9928 | 3284 | 16572 |
| Delhi | 13840 | 3386 | 24294 | 1384 | 339 | 288 | 692 | 169 | 144 | 11764 | 2878 | 2446 |
| Gujarat | 10716 | 3213 | 18219 | 1072 | 321 | 1822 | 536 | 161 | 911 | 9109 | 2731 | 15486 |
| Haryana | 1589 | 0 | 4618 | 159 | 0 | 462 | 79 | 0 | 231 | 1350 | 0 | 3925 |
| Jammu & Kashmir | 2372 | 1040 | 3705 | 237 | 104 | 370 | 119 | 52 | 185 | 2016 | 884 | 3149 |
| Karnataka | 6121 | 2820 | 9422 | 612 | 282 | 942 | 306 | 141 | 471 | 5203 | 2397 | 8008 |
| Kerala | 1362 | 0 | 3035 | 136 | 0 | 304 | 68 | 0 | 152 | 1158 | 0 | 2580 |
| Madhya Pradesh | 6487 | 0 | 13379 | 649 | 0 | 1338 | 324 | 0 | 669 | 5514 | 0 | 11372 |
| Maharashtra | 91428 | 67487 | 115369 | 9143 | 6749 | 11537 | 4571 | 3374 | 5768 | 77714 | 57364 | 98064 |
| Odisha | 1627 | 0 | 3573 | 163 | 0 | 357 | 81 | 0 | 179 | 1383 | 0 | 3037 |
| Punjab | 115 | 0 | 2967 | 12 | 0 | 297 | 6 | 0 | 148 | 98 | 0 | 2522 |
| Rajasthan | 8962 | 3571 | 14353 | 896 | 357 | 1435 | 448 | 179 | 718 | 7618 | 3036 | 12200 |
| Tamil Nadu | 9163 | 0 | 35097 | 916 | 0 | 351 | 458 | 0 | 1755 | 7789 | 0 | 29833 |
| Telangana | 1854 | 0 | 4393 | 185 | 0 | 439 | 93 | 0 | 220 | 1576 | 0 | 3734 |
| Uttar Pradesh | 6716 | 936 | 12496 | 672 | 94 | 1250 | 336 | 47 | 625 | 5709 | 796 | 10621 |
| West Bengal | 4157 | 2361 | 5954 | 416 | 236 | 595 | 208 | 118 | 298 | 3534 | 2007 | 5061 |

Source: Based on the author's calculations using R-studio

**Table 6:** Forecast of Active Cases of COVID-19 and Required Health Infrastructure for India and its States on 15th July, 2020, India

| STATES | Active Cases | 95% CI |  | ICU | 95% CI |  | Ventilators | 95% CI |  | Beds | 95% CI |  |
| --- | --- | --- | --- | --- | --- | --- | --- | --- | --- | --- | --- | --- |
|  | PE | LB | UB | PE | LB | UB | PE | LB | UB | PE | LB | UB |
| India | 296472 | 196820 | 396125 | 29647 | 19682 | 39612 | 14824 | 9841 | 19806 | 252001 | 167297 | 336706 |
| Andhra Pradesh | 1131 | 0 | 4685 | 113 | 0 | 468 | 57 | 0 | 234 | 961 | 0 | 3982 |
| Bihar | 17479 | 2997 | 31961 | 1748 | 300 | 3196 | 874 | 150 | 1598 | 14857 | 2547 | 27167 |
| Delhi | 16773 | 142 | 33404 | 1677 | 14 | 12 | 839 | 7 | 6 | 14257 | 121 | 102 |
| Gujarat | 12299 | 572 | 24027 | 1230 | 57 | 2403 | 615 | 29 | 1201 | 10455 | 486 | 20423 |
| Haryana | 2200 | 0 | 7351 | 220 | 0 | 735 | 110 | 0 | 368 | 1870 | 0 | 6248 |
| Jammu & Kashmir | 2986 | 867 | 5105 | 299 | 87 | 510 | 149 | 43 | 255 | 2538 | 737 | 4339 |
| Karnataka | 8039 | 2707 | 13372 | 804 | 271 | 1337 | 402 | 135 | 669 | 6833 | 2301 | 11366 |
| Kerala | 1807 | 0 | 4520 | 181 | 0 | 452 | 90 | 0 | 226 | 1536 | 0 | 3842 |
| Madhya Pradesh | 7864 | 0 | 18866 | 786 | 0 | 1887 | 393 | 0 | 943 | 6685 | 0 | 16036 |
| Maharashtra | 114873 | 76308 | 153438 | 11487 | 7631 | 15344 | 5744 | 3815 | 7672 | 97642 | 64862 | 130422 |
| Odisha | 1969 | 0 | 5116 | 197 | 0 | 512 | 98 | 0 | 256 | 1674 | 0 | 4349 |
| Punjab | 115 | 0 | 3502 | 12 | 0 | 350 | 6 | 0 | 175 | 98 | 0 | 2977 |
| Rajasthan | 11338 | 2703 | 19972 | 1134 | 270 | 1997 | 567 | 135 | 999 | 9637 | 2297 | 16977 |
| Tamil Nadu | 9793 | 0 | 51497 | 979 | 0 | 515 | 490 | 0 | 2575 | 8324 | 0 | 43772 |
| Telangana | 2310 | 0 | 6339 | 231 | 0 | 634 | 115 | 0 | 317 | 1963 | 0 | 5388 |
| Uttar Pradesh | 8426 | 0 | 17660 | 843 | 0 | 1766 | 421 | 0 | 883 | 7162 | 0 | 15011 |
| West Bengal | 5041 | 2275 | 7807 | 504 | 228 | 781 | 252 | 114 | 390 | 4285 | 1934 | 6636 |

### 3.6 Estimates of required healthcare infrastructure at state-level

**Table 4, 5, and 6** show the required number of isolation beds, ICU beds and ventilators for patients that will be suffering from COVID-19 on 15^th^, 30th June and 15th July respectively for Indian states. The overall picture for estimates shows a huge variation across the states. By 15th June, Kerala, Haryana, Punjab and Andhra Pradesh will recover most of the active cases and requires less than 1000 isolation beds, around 100 ICU beds and 50 ventilator beds according to our point estimates. While, on the other hand, Maharashtra, with the largest number of active cases shows an alarming situation and will require 57786 isolation beds (95% CI: 47708, 67864), 6798 ICU beds (95% CI: 5613, 7984) and 3399 ventilator beds (95% CI: 2806, 3992).

At the end of June month, Kerala, Haryana, Punjab, Odisha, Telangana and Andhra Pradesh will require less than 2000 isolation beds, under 200 ICU beds and less than 100 ventilator beds. While, in Maharashtra, will require 77714 isolation beds (95% CI: 57364, 98064), 9143 ICU beds (95% CI: 6749, 11537) and 4571 ventilator beds (95% CI: 3374, 5768).

At the mid of July, Kerala, Haryana, Punjab, Odisha, Telangana and Andhra Pradesh will require less than 2000 isolation beds, around 200 ICU beds and 100 ventilator beds. While, on the other hand, Maharashtra, with the largest number of active cases and will require 97642 isolation beds (95% CI: 64862, 130422), 11487 ICU beds (95% CI: 7631, 15344) and 5744 ventilators (95% CI: 3815, 7672) will be required by 15th July.

## 4. Discussion and Conclusion

Our current forecasts for confirmed and active cases are in line with the actual number of cases. On 2nd June, India has 207,186 confirmed cases, while our forecasts suggested 206,764 confirmed cases (95% CI: 194194, 219334). India had 101,070 active cases on 2nd June, while our forecasts suggested that India will have 104,526 active cases (95% CI: 96145, 112907). Our results also show that daily confirmed cases will increase at a faster pace as around mid-July India will be getting around 16,000-17,000 daily confirmed cases. By 15th July, we forecast that the total confirmed cases will be around 7.5 lakhs, whereas total active cases will be close to 2.96 lakhs based on our point estimates for India. Maharashtra will be the most affected state even at the 15th July with 1.9 Lakh confirmed cases and 1.15 Lakh active cases while Punjab will be least affected having 2648 confirmed cases and only 115 active cases based on our point estimates. The total number of isolation beds required for India is 2,52,001 (95% CI: 167297, 336706). Our estimates also show that 29,647 ICU beds (95% CI: 19682, 39612) and 14,824 ventilators (95% CI: 9841, 19806) will be required in India by the 15th July. When it comes to states, Maharashtra will be the most affected state and will require 97,642 isolation beds (95% CI: 64862, 130422).

On 3rd June, with respect to the health infrastructure in the country for the management of COVID-19, 952 dedicated COVID hospitals with 1,66,332 Isolation beds, 21,393 ICU beds and 72,762 Oxygen supported beds are available. 2,391 dedicated COVID Health Centres with 1,34,945 Isolation beds; 11,027 ICU beds and 46,875 Oxygen supported beds have also been operationalized. Based on the current preparedness of the Indian healthcare infrastructure, it seems likely that India will not be facing a shortage at the national level. However, the health infrastructure stress will likely be acute in high-burden states, led by Maharashtra, Tamil Nadu and Delhi as they have more than 10,000 active cases on 2nd June. In particular, Maharashtra is the most affected state with around 40,000 active cases as on 2nd June. On 29th May, BrihanMumbai Municipal Corporation (BMC) reported that almost all intensive care unit (ICU) beds for COVID-19 patients were occupied in Mumbai. They also tell that the city is already using 72% of the total 373 ventilators available for COVID-19 treatment (Mint, 2020). This shows an alarming situation for Maharashtra and in particular, Mumbai. The most affected state Maharashtra needs to prepare at least 1 lakh isolation beds, 12,000 ICU beds and 6000 ventilators by 15th July to fight this pandemic efficiently.

Our forecasts are based on the data which grossly under-reported the cases as found by many studies for India. Rao *etal*, (2020), projected that India might be detecting 1 out of 4 cases of COVID-19 based on his mathematical model. Another study by (Goli and James, 2020) also find that India is detecting just 3.6% of the total number of infections of COVID-19 with a huge variation across its states. They also suggest that India must increase its testing capacity and go for widespread testing to know the real picture of the pandemic. Lack of coverage of testing is not revealing the true prevalence of COVID-19. (Subramanian and James, 2020) suggested that for detection of the true prevalence of COVID-19 infections in the country, India can adopt the well-established National Family Health Survey (NFHS) framework as a solution to ascertain the true prevalence of COVID-19. Understanding the urgency of the situation, on 8th May, The Indian Council of Medical Research (ICMR) announced that it will conduct a study in 75 affected districts across the country to identify people who were exposed to the novel coronavirus infection and yet showed mild or no symptoms. The study can help ascertain whether there has been community transmission of the respiratory disease in those areas or not. This much-needed study by ICMR will reveal the true prevalence of asymptomatic COVID-19 infected people to check for community spread (The Economic Times, 2020). The data issues of COVID-19 such as under-reporting, lack of coverage etc. results in an underestimation of the cases, which in turn will lead to underestimation of the forecasts in our study.

Our forecasts show an alarming situation for India in the future. In coming months, the actual numbers can go higher than our forecasts of confirmed cases, active cases and healthcare infrastructure as migrant workers are returning home in large numbers currently in large numbers (Tyagi *et al*, 2020). Currently, India has lifted lockdown with restrictions applying only on the containment zones. This will lead to a surge in the number of daily confirmed and active cases. The requirement of isolation beds, ICUs and ventilators will also be increased in that scenario. So, India and its majorly affected states like Maharashtra, Gujarat, Tamil Nadu, Bihar and Delhi need to be well prepared for the pandemic challenge in future and focus on increasing their healthcare infrastructure, and other states should also remain alert till the pandemic completely recedes.

## Data Availability

The daily confirmed and active cases data used in this study are easily available on www.covid19india.org and updated on a daily basis.

## APPENDIX

**Fig. 4.**
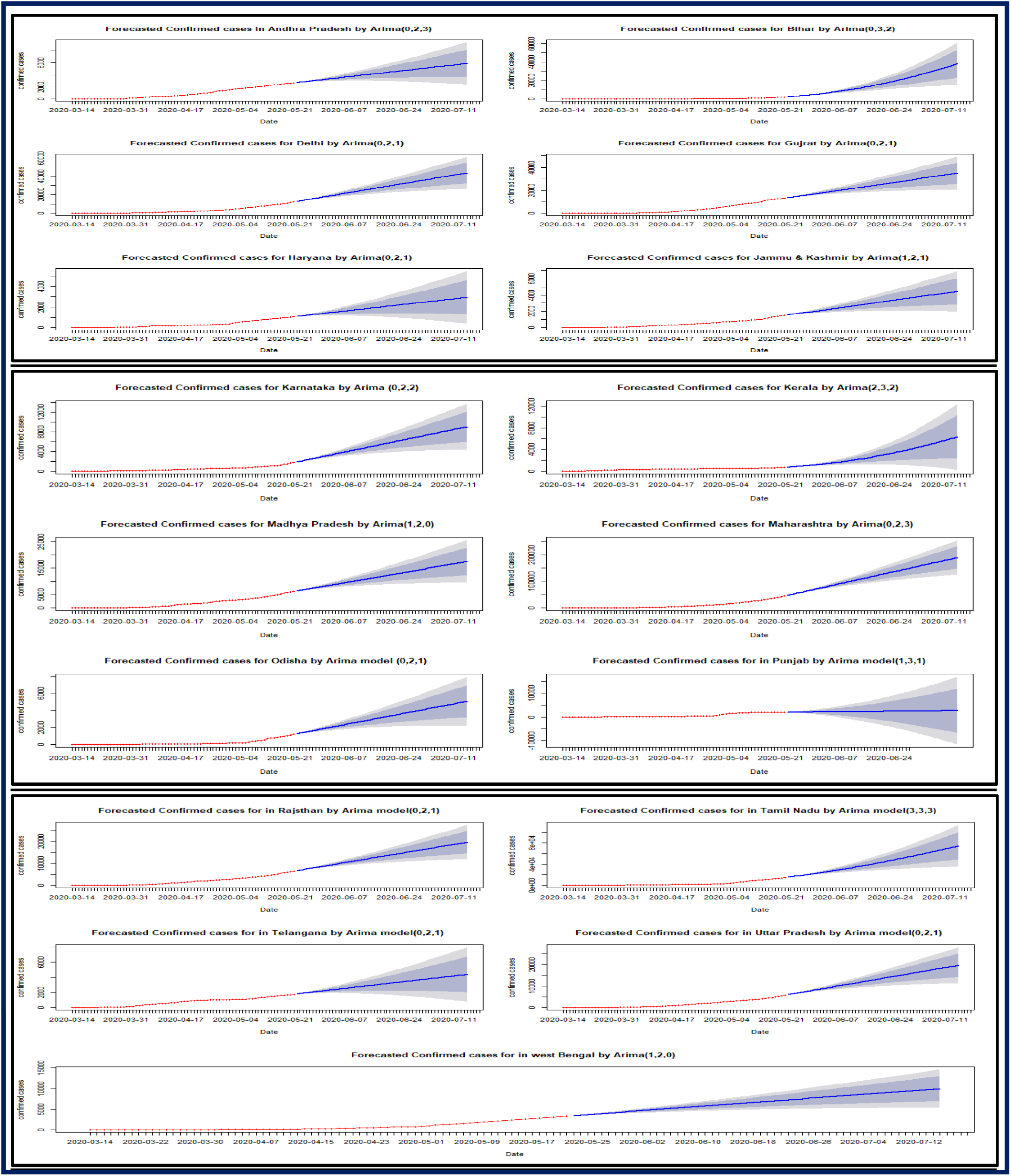
Forecast trend of Confirmed cases of COVID-19 for Indian States

**Fig. 4.**
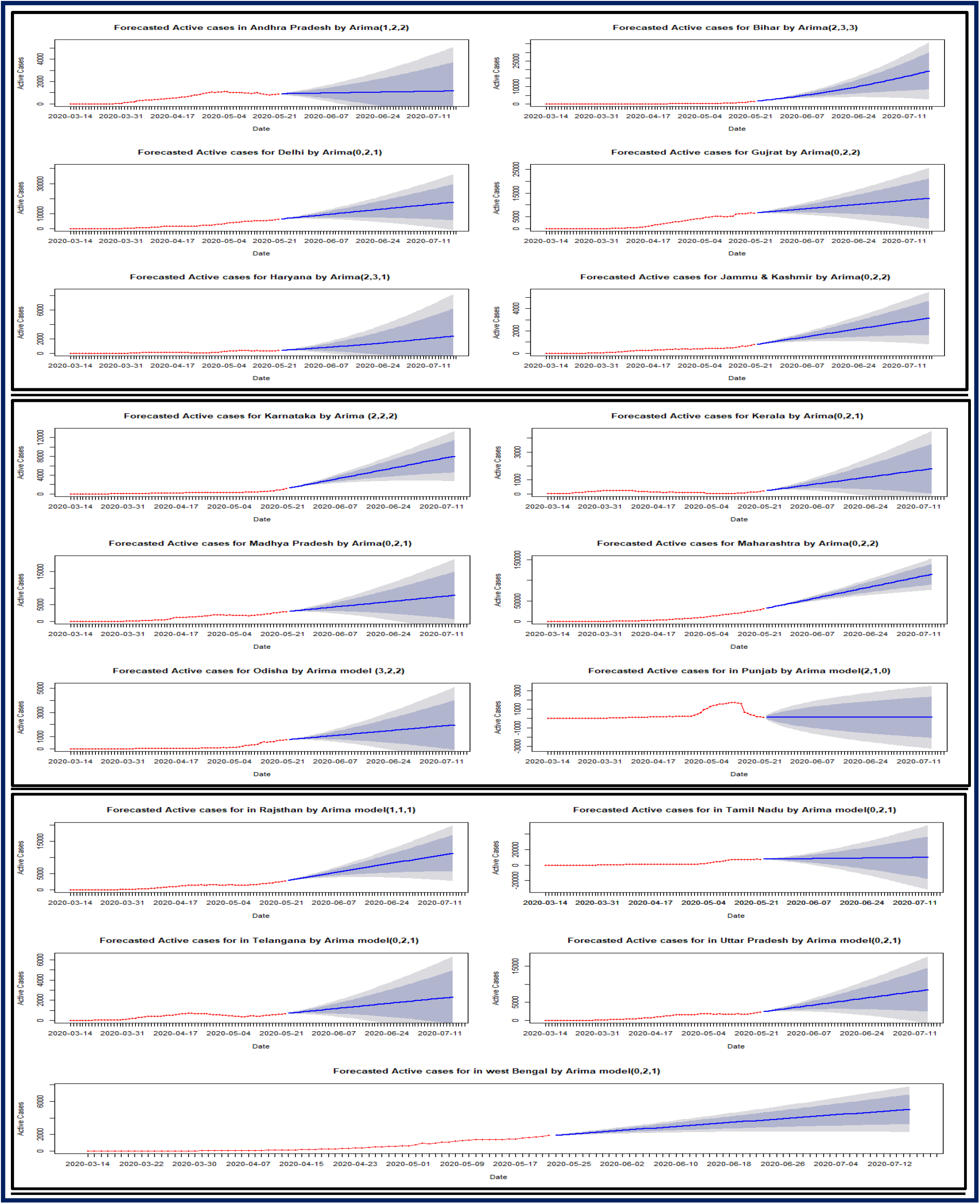
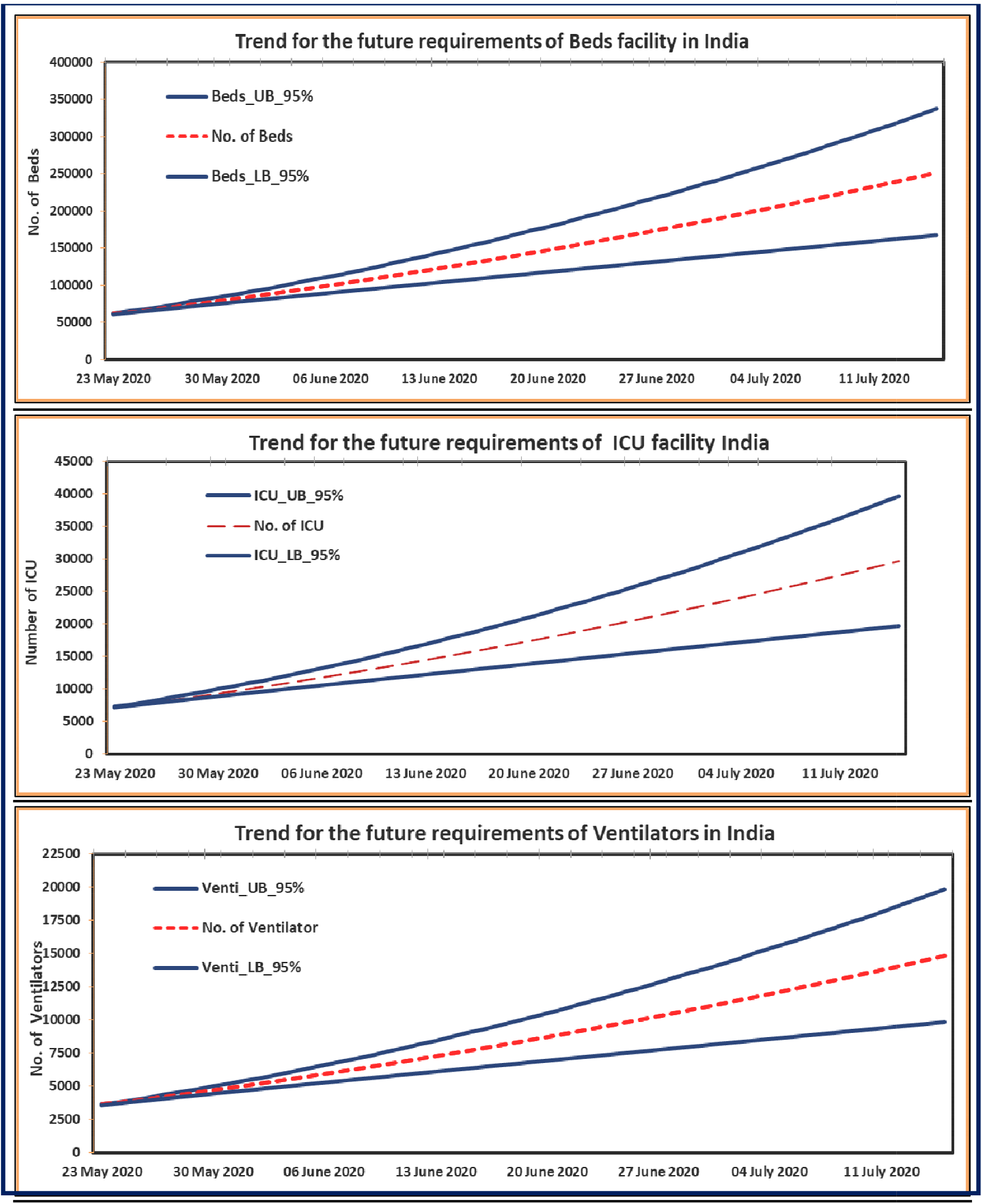
Forecast trend of Active cases of COVID-19 for Indian States

## References

1. Chakraborty, T., & Ghosh, I. (2020). Real-time forecasts and risk assessment of novel coronavirus (COVID-19) cases: A data-driven analysis. Chaos, solitons, and fractals, 135, 109850. Advance online publication. https://doi.org/10.1016/j.chaos.2020.109850

2. Goli, S and James, K.S. (2020) How much India detecting SARS-CoV-2 Infections? A model-based estimation. medRxiv preprint doi: https://doi.org/10.1101/2020.04.09.20059014.

3. Guan, W. J., Ni, Z. Y., Hu, Y., Liang, W. H., Ou, C. Q., He, J. X., … & Du, B. (2020). Clinical characteristics of coronavirus disease 2019 in China. New England journal of medicine, 382(18), 1708–1720.

4. Gupta R., Pal S. K. (2020), Trend Analysis and Forecasting of COVID-19 outbreak in India. MedRxiv. https://doi.org/10.1101/2020.03.26.200445110

5. Kumar P., Kalita H., Patairiya S., Sharma Y. D., Nanda C., Rani M., Rahmai J., Bhagavathula A. S. (2020), Forecasting the dynamics of COVID-19 Pandemic in Top 15 countries in April 2020 through ARIMA Model with Machine Learning Approach. MedRxiv. https://doi.org/10.1101/2020.03.30.20046227

6. Perone, G. (2020). An ARIMA model to forecast the spread and the final size of COVID-2019 epidemic in Italy. *medRxiv.*

7. Rao, A. S., Krantz, S. G., Kurien, T., Bhat, R., & Kurapati, S. (2020). Model-based retrospective estimates for covid-19 or Coronavirus in India: continued efforts required to contain the virus Spread. Current Science, 118(7), 1023–1025.

8. Remuzzi, A., & Remuzzi, G. (2020). COVID-19 and Italy: what next? The Lancet.

9. Singh, R. K., Rani, M., Bhagavathula, A. S., Sah, R., Rodriguez-Morales, A. J., Kalita, H., Nanda, C., Sharma, S., Sharma, Y. D., Rabaan, A. A., Rahmani, J., & Kumar, P. (2020). Prediction of the COVID-19 Pandemic for the Top 15 Affected Countries: Advanced Autoregressive Integrated Moving Average (ARIMA) Model. JMIR public health and surveillance, 6(2), e19115. https://doi.org/10.2196/19115

10. Subramanian, S. V., & James, K. S. (2020). Use of the Demographic and Health Survey framework as a population surveillance strategy for COVID-19. The Lancet Global Health.

11. Tandon, H., Ranjan, P., Chakraborty, T., & Suhag, V. (2020). Coronavirus (COVID-19): ARIMA based time-series analysis to forecast near future. arXiv preprint arXiv:2004.07859.

12. Tiwari, S., Kumar, S., & Guleria, K. (2020). Outbreak Trends of Coronavirus Disease-2019 in India: A Prediction. Disaster medicine and public health preparedness, 1–6. Advance online publication. https://doi.org/10.1017/dmp.2020.115

13. Tyagi, R., Dwivedi, L.K., & Sanzgiri, A. (2020). Estimation of Effective Reproduction Numbers for COVID-19 using Real-Time Bayesian Method for India and its States. Paper: 13 IIPS Analytical Series on COVID-19

14. World Health Organization. (2020). Coronavirus disease 2019 (COVID-19): situation report, 94. **Web-based Sources**

15. Forecasting COVID-19 cases in India https://towardsdatascience.com/forecasting-CQVID-19-cases-in-india-c1c410cfc730

16. India Today (2020). Kerala reports first confirmed coronavirus case in India. https://www.indiatoday.in/india/story/kerala-reports-first-confirmed-novel-coronavirus-case-in-india-1641593-2020-01-30

17. Livemint (2020). Mumbai coronavirus update: 99% ICU beds in hospitals are occupied, says BMC. https://www.livemint.com/news/india/coronavirus-in-mumbai-99-icu-beds-in-hospitals-is-occupied-says-bmc-11590742717577.html

18. Press Information Bureau (2020). ‘Government of India issues Orders prescribing lockdown for containment of COVID-19 Epidemic in the country’, 24th March.

19. Press Information Bureau (2020). ‘PIB’S DAILY BULLETIN ON COVID-19’, 8th May.

20. Press Information Bureau (2020). ‘PIB’S DAILY BULLETIN ON COVID-19’, 3rd June.

21. The Economic Times (2020). ICMR to study prevalence of asymptomatic COVID-19 infected people to check for community spread. Retrieved from https://economictimes.indiatimes.com/news/politics-and-nation/icmr-to-study-prevalence-of-asymptomatic-covid-19-infected-people-to-check-for-community-spread/articleshow/75631365.cms?from=mdr

